# Measuring autistic traits in Hungarian adults: Psychometric evaluation of the revised Hungarian Autism Spectrum Quotient (AQ-50-HU-R)

**DOI:** 10.64898/2026.09.01.26361970

**Authors:** Dániel Sörnyei, Fanni Mercédesz Kovács, Tünde Benedek, Dorottya Őri, Kinga Farkas

**Affiliations:** Department of Clinical Psychology, Semmelweis University, Budapest, Hungary; Department of Psychiatry and Psychotherapy, Semmelweis University, Budapest, Hungary; Department of Psychiatry A, Nyírő Gyula National Institute of Psychiatry and Addictology, Budapest, Hungary; Institute of Behavioral Sciences, Semmelweis University, Budapest, Hungary; Department of Mental Health, Heim Pál National Pediatric Institute, Budapest, Hungary

## Abstract

The Autism Spectrum Quotient (AQ-50) is widely used to assess autistic traits, yet its Hungarian version has not been psychometrically evaluated. We assessed the reliability, factor structure, temporal stability, convergent validity, and clinical utility of the Hungarian AQ-50 and a revised translation (AQ-50-HU-R) in two samples (*N_1_* = 1967; *N_2_* = 423), including autistic and non-autistic participants. The AQ-50-HU-R showed high internal consistency and test-retest reliability. A bifactor model provided the best fit (*χ^2^*[1125] = 1650.433, *p* < 0.001; CFI = 0.991; TLI = 0.990; RMSEA = 0.033 [90% CI = 0.030-0.037]; SRMR = 0.083), with 71% of common variance attributable to a general autistic traits factor. The total score distinguished clinically verified autistic participants from participants reporting no ASD diagnosis (AUC = 0.906), with a cutoff of 25. Associations with ADOS scores were weak or nonsignificant. The AQ-50-HU-R is best interpreted as a reliable total-score screening measure, supporting referral for comprehensive autism assessment.

## Introduction

Autism is an extensively studied neurodevelopmental spectrum condition defined by persistent differences in social communication and interaction, as well as by restricted, repetitive patterns of behavior, interests, or activities [1]. Although the core features of autism typically emerge in the early developmental period, they persist into adulthood and are often accompanied by co-occurring psychiatric conditions, including anxiety, depression, sleep-wake disturbances, attention-deficit/hyperactivity disorder (ADHD), and obsessive-compulsive disorder (OCD) [2,3]. The global prevalence of autism spectrum disorder (ASD) in formal diagnostic manuals is traditionally estimated at approximately 1% in the general population, with an overall male-to-female ratio of around 3:1 [1,4]. However, epidemiological studies reported substantially higher prevalence rates both among children [5,6] and within adult psychiatric populations. For instance, in the United Kingdom, autism was identified in an estimated 4.8% of individuals receiving psychiatric care [7]. Similarly, a study conducted in Sweden observed that the prevalence of autism among adults in outpatient psychiatric services was at least 18.9%, with an additional 5-10% exhibiting subclinical features [8].

Despite elevated prevalence rates, autism often remains undetected in routine psychiatric assessments, suggesting a substantial risk of underdiagnosis in everyday clinical practice. This concern is best exemplified by findings suggesting that between 59% and 72% of autistic adults remain undiagnosed [9]. In parallel, increasing attention is being directed toward the female autism phenotype, raising the possibility that underdiagnosis among females may reflect differences in the presentation of autistic traits compared to traditional diagnostic criteria, including greater camouflaging [4,10]. Early identification of autism facilitates timely access to psychosocial support services, which have been associated with improvements in social functioning, psychological well-being, and overall quality of life [11,12].

The diagnostic process for ASD involves integrating multiple sources of information across developmental stages, including clinical interviews with the individual and their caregivers, direct behavioral observations, review of historical medical records, and educational or psychological reports, typically gathered by a multidisciplinary team [13,14]. Cross-sectional diagnostic instruments, often referred to as “gold-standard” tools, include the *Autism Diagnostic Observation Schedule* (ADOS [15,16]) – a semi-structured behavioral assessment administered directly to the individual – and the *Autism Diagnostic Interview-Revised* (ADI-R [17]), a semi-structured interview conducted with caregivers. While these instruments offer robust diagnostic validity, they are also resource-intensive and time-consuming. This highlights the need for efficient, sensitive screening tools that can guide clinicians in determining whether a comprehensive diagnostic evaluation is warranted. Commonly used screening measures include the *Ritvo Autism Asperger Diagnostic Scale – Revised* (RAADS-R [18]), the *Social Responsiveness Scale, Second Edition* (SRS-2 [19]), and the *Autism Spectrum Quotient* (AQ-50 [20]).

The AQ-50 [20] was one of the earliest self-report questionnaires developed to screen for autistic traits in adults with average intellectual functioning, as well as to assist in the diagnosis of ASD. Consistent with the broad autism spectrum perspective [21–23], which conceptualizes autistic characteristics as dimensional traits, the original version of AQ-50 assesses five theoretically derived domains: *social skill, attention switching, attention to detail, communication,* and *imagination.* However, subsequent research has consistently failed to replicate the original five-factor structure. Importantly, English et al. noted considerable inconsistencies across international studies regarding the scoring format (binary vs. Likert scale), the item composition of factors, the number of empirically derived factors, and appropriate clinical cutoff scores [24]. Another interesting observation is that while factors reflecting social differences and restricted interests showed relative stability, those related to imagination were inconsistently defined across studies [24–27]. These inconsistencies have inspired competing theories, including proposals that autistic traits are best conceptualized as a largely unitary construct, as well as models that emphasize a set of partially distinct features, with individuals showing distinct profiles across these trait domains [24,28,29].

In terms of clinical utility, the seminal study by Baron-Cohen et al. [20] proposed a cutoff score of 32 out of 50 points to indicate high levels of autistic traits. Subsequent studies across diverse countries and populations suggested alternative, typically lower thresholds for distinguishing autistic individuals from neurotypical populations [30–33]. Taken together, these findings underscore the variability and lack of consensus regarding the psychometric properties of AQ-50. Nevertheless, it is increasingly recognized that, when applied with caution, the AQ-50 can serve as a useful screening tool for identifying individuals who may benefit from further evaluation and support related to autistic traits [23].

Although Hungarian translations of the AQ-50 are currently used in both research and clinical contexts, to the best of our knowledge, no study to date has evaluated the reliability and validity of the Hungarian version. To address this issue, the present study aims to refine the translation and improve the instrument’s linguistic accuracy, and to examine the psychometric properties and clinical applicability of the Hungarian version of the AQ-50. Our broader goal is to provide a psychometrically sound self-report tool for assessing autistic traits in Hungarian-speaking populations. Based on previous international research, we do not assume that the original five-factor structure will replicate in Hungarian samples. Therefore, our objective is to test several competing models empirically, and we expect that a revised Hungarian version of the AQ-50 will demonstrate acceptable reliability and a satisfactory factor structure, thereby supporting its use as a clinically relevant screening instrument. Thus, this study has the potential to advance autism research in Hungary, contribute to the existing literature, and support everyday mental health practice. Specifically, it may help improve the screening of adults who may benefit from a more comprehensive evaluation for autism.

## Materials and Methods

### Procedure

The study was conducted in Hungarian and followed a cross-sectional design with a longitudinal retest component. Sample sizes were not determined using an a priori power analysis. Instead, they comprised all eligible participants retained through convenience recruitment, as described below. We report all data exclusions and all measures analyzed in the study. No experimental manipulations were conducted.

#### Procedure for Sample 1

Participants in Sample 1 were recruited between 1 November 2021 and 30 April 2022. Recruitment for the clinically verified autistic subsample subsequently continued from 1 January 2022 to 30 April 2024. In the first phase, we recruited participants through multiple channels, including social media platforms and direct outreach to outpatient services. We recruited autistic individuals from the Outpatient Unit of the Department of Psychiatry and Psychotherapy at Semmelweis University and through organizations supporting autistic individuals. Potential participants could contact the research team by email or access the online questionnaire battery directly via a link or QR code. We provided a dedicated email address for questions and additional support. Participants completed the self-report questionnaires on the formr.org platform [34], which collected demographic and clinical information, as well as responses to the AQ-50.

After completing the online survey, participants who reported a medical diagnosis of ASD were invited to provide their contact details to take part in the second phase of data collection, which formed part of a larger study examining perceptual, social-cognitive, and neuroimaging characteristics in individuals on the autism and schizophrenia spectrum, as well as neurotypical participants. In this phase, trained psychiatrists administered the ADOS to quantify behavioral features of autism. Participants were also asked to submit medical documentation confirming their ASD diagnosis, which a psychiatrist subsequently validated. In the third phase, we evaluated the test-retest reliability of the AQ-50-HU by having all participants complete the online questionnaire battery again after a 6-month interval.

#### Procedure for Sample 2

Participants in Sample 2 were recruited between 5 August 2025 and 30 June 2026. Following linguistic accuracy updates to the original Hungarian version (described in detail in the *Measures* section), we recruited a second sample to complete both the original translation and the revised items (AQ-50-HU-R) using the same recruitment procedures. Test-retest reliability was evaluated by asking participants in Sample 2 to complete the online questionnaire battery again after a 1-month interval.

The study procedure complied with the ethical standards of national and institutional committees on human experimentation and with the Declaration of Helsinki. Ethical approval was granted by the Semmelweis University Regional and Institutional Committee of Science and Research Ethics and the Medical Research Council (approval numbers: RKEB 158/2021; RKEB 159/2021 and BM/14269-3/2025). Before data collection, all participants provided electronic written informed consent.

### Participants

#### Sample 1

A total of 2008 participants agreed to take part in the first survey. As described in the *Procedure* section, we recruited participants through social media platforms, outpatient services, and organizations supporting autistic individuals. To ensure data quality, only individuals aged 18 to 65 years were included. Additionally, we required all participants to have completed at least 8 years of elementary education, and excluded those reporting more than 30 years. Participants with a diagnosis of a schizophrenia spectrum disorder were also excluded because of the substantial overlap between negative symptoms and autistic traits. Sample 1 consisted of 1967 participants, with 671 (34.11%) identifying as men, 1237 (62.89%) as women, and 59 (3.00%) as another gender. A total of 274 (13.93%) individuals self-reported a diagnosis of ASD, of which 40 participants (2.03%) were verified through medical documentation during the second phase of data collection. The remaining 1693 participants (86.07%) reported having no ASD diagnosis and were therefore classified as non-ASD. The verified ASD subsample (*N =* 40), which participated in the second, clinical phase of data collection, included 24 (60%) men, 12 (30%) women, and 4 (10%) individuals identifying as another gender. The retest subsample (*N =* 325) comprised 97 (29.85%) men, 217 (66.77%) women, and 11 (3.38%) individuals of other genders.

#### Sample 2

A total of 492 participants started the second survey. Data collection and data filtering procedures were identical to those described above. Sample 2 comprised 423 participants, of whom 113 (26.7%) identified as men, 298 (70.5%) as women, and 12 (2.8%) as another gender. Of these, 140 (33.1%) participants self-reported an ASD diagnosis, and 283 (66.9%) were classified as non-ASD. The verified ASD subsample (*N =* 44) included 15 (34.1%) men, 25 (56.8%) women, and 4 (9.1%) individuals identifying as another gender. The retest subsample (*N* = 97) included 18 (18.6%) men, 75 (77.3%) women, and 4 (4.1%) participants identifying as another gender. A detailed breakdown of demographics is provided in Table 1. We obtained electronic written informed consent from all participants before the online surveys began.

**Table 1.**
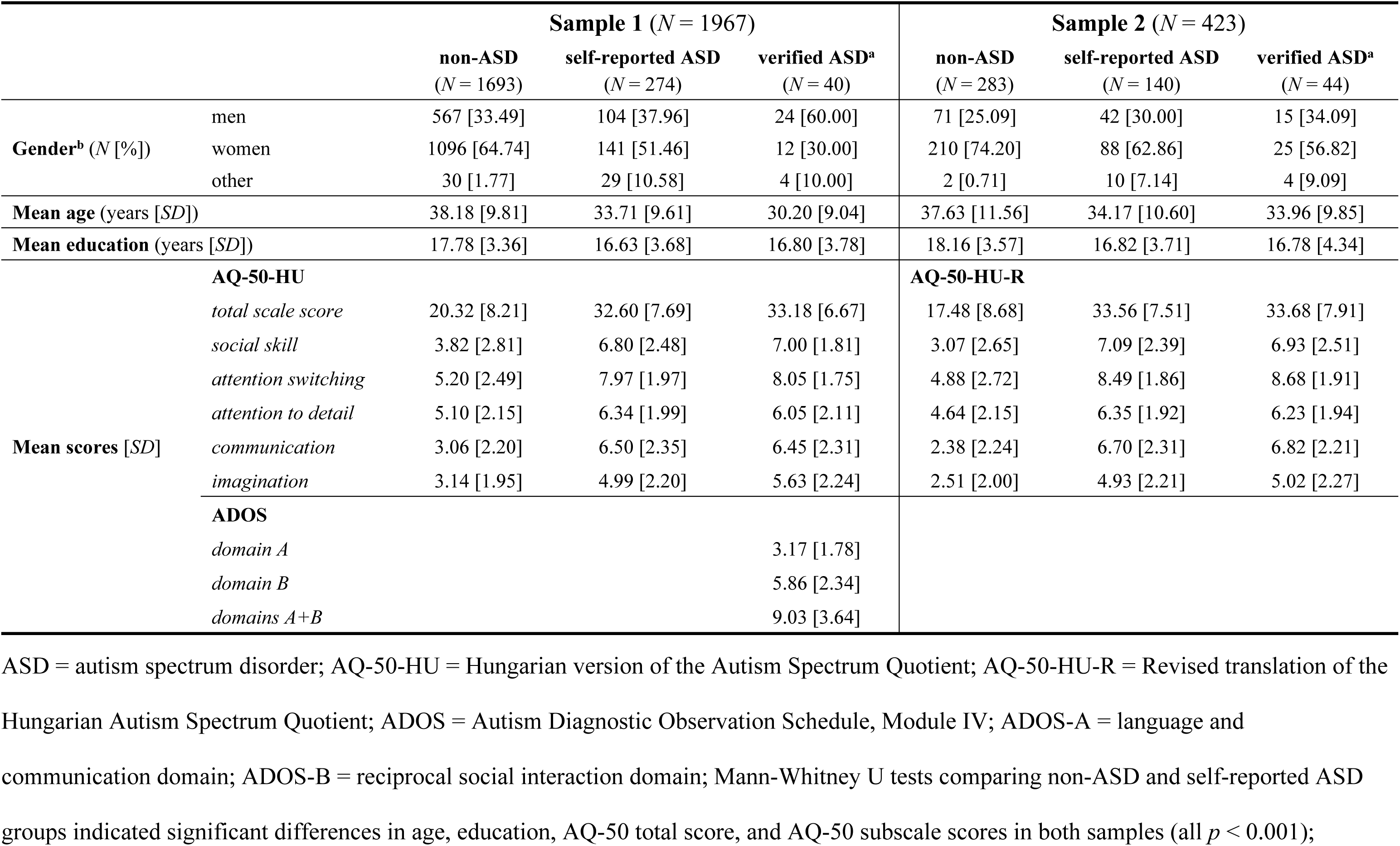
Demographics and scale score distributions across samples and groups.

### Measures

#### Autism Spectrum Quotient (AQ-50)

We assessed autistic traits using the *Autism Spectrum Quotient* (AQ-50 [20]). This self-report questionnaire was originally designed as a screening tool for adults with average intellectual functioning. The AQ-50 evaluates the extent of autistic characteristics across five distinct domains: *social skill, attention switching, attention to detail, communication,* and *imagination.* Each domain comprises 10 items, and participants have to indicate their responses on a 4-point Likert scale (*definitely agree; slightly agree; slightly disagree; definitely disagree*). Scoring can be performed in two ways: using the original binary method (e.g., for non-reversed items, *definitely agree* and *slightly agree* = 1 point; *slightly disagree* and *definitely disagree* = 0 points) or using the 4-point Likert scale to capture greater response variability [24,35,36]. In the present study, we employed the binary scoring method. Although an official Hungarian adaptation of the questionnaire has not yet been developed, three slightly different unofficial translations have been used in Hungarian research and clinical practice for several years. To ensure comparability with previous research, we selected the most widely available version for Sample 1. During data collection, however, we identified minor linguistic inaccuracies in several items. Although these issues did not fundamentally alter the intended content, the wording did not fully capture the nuance of the original items. To address this, the research team identified the affected items and developed a revised translation using an iterative back-translation procedure conducted under the supervision of the original authors. The revised wording was developed by clinicians with experience working with autistic individuals, all of whom had advanced English proficiency. As a result, we revised seven items (1, 9, 11, 38, 41, 48, and 50) to improve linguistic accuracy. We then conducted a pilot comprehension test and a review of the full questionnaire in two focus groups: one with autistic participants (*N* = 9) and one with non-autistic participants (*N* = 8). Final item wording was determined by integrating their feedback with the translation team’s recommendations. The revised Hungarian version (AQ-50-HU-R), including the scoring key, is available from the authors upon request.

#### Autism Diagnostic Observation Schedule, Module IV (ADOS)

To assess differences in communication and reciprocal social interaction, we used the *Autism Diagnostic Observation Schedule, Module IV* (ADOS [15,16]). The ADOS was included to evaluate potential discrepancies between self-reported autistic traits and clinician-ascertained behavioral features, and to provide evidence for the convergent validity of the AQ-50-HU. The ADOS is a semi-structured behavioral assessment administered by a trained clinician and typically requires 45-60 minutes to complete. It is widely regarded as the “gold standard” instrument for ASD diagnosis and comprises five domains: (A) *language and communication,* (B) *reciprocal social interaction,* (C) *imagination,* (D) *stereotyped behaviors and restricted interests,* and (E) *other abnormal behaviors.* The diagnostic algorithm defines a cutoff score for ASD based on domains A and B. Accordingly, our study focused exclusively on these two domains to assess differences in language, communication, and reciprocal social interaction. The ADOS Manual [16] reports high composite reliability for these domains (Cronbach’s α = 0.91-0.94). In contrast, we observed lower internal consistency estimates in the present study (Cronbach’s α = 0.66; McDonald’s ω = 0.62), which may reflect the relatively small size and restricted range of the clinical subsample.

### Statistical analyses

We conducted statistical analyses and visualization in JASP 0.19.3.0 [37], R [38], and RStudio [39] using the packages *dplyr* [40], *BifactorIndicesCalculator* [41], *gghalves* [42], *ggplot2* [43], *lavaan* [44], *pROC* [45], *patchwork* [46], *semTools* [47], and *tidyverse* [48]. Statistical significance was set at *p* < 0.05. We assessed normality using the Shapiro-Wilk test. Internal consistency of scale scores was evaluated with Cronbach’s alpha (≥ 0.70 acceptable) and McDonald’s omega (≥ 0.70 acceptable) coefficients.

To examine the factorial structure of the AQ-50, we conducted confirmatory factor analyses (CFA). Consistent with the conceptualization of autistic traits as continuously distributed across the population [23], we performed CFAs in the full samples (Sample 1: *N* = 1967; Sample 2: *N* = 423). Factor scaling was based on factor variances. Missing data were handled via pairwise deletion. Since the items represent ordered categorical responses, we estimated models using diagonally weighted least squares (DWLS). Model fit was evaluated using the chi-square (*χ^2^*), Root Mean Square Error of Approximation (RMSEA; values ≤ 0.07 indicating acceptable fit), Standardized Root Mean Square Residual (SRMR; ≤ 0.08), Comparative Fit Index (CFI; ≥ 0.90), and Tucker-Lewis Index (TLI; ≥ 0.90). We evaluated composite factor reliability using omega and hierarchical omega indices (ω and ω_H_ > 0.50), and computed Hancock’s H coefficient (H > 0.70) to assess construct replicability. For bifactor models, we also computed explained common variance (ECV) to compare the relative contribution of general versus specific factors. Based on prior research [20,24,29], we compared several competing models in Sample 1: (1) the original 5-factor model; (2) a unidimensional (single-factor) model of autistic traits, and (3) a bifactor model representing one general factor and five specific domain factors. In Sample 2, we restricted analyses to the bifactor model.

We evaluated sensitivity and specificity for discriminating between ASD and non-ASD participants using receiver operating characteristic (ROC) analyses. Test-retest reliability was examined by calculating Spearman’s rank correlations (*r_s_*) between AQ-50 scores obtained at two time points. We examined convergent validity using Spearman’s correlations (*r_s_*) between AQ-50 total and subscale scores and ADOS total and domain scores in the diagnostically verified ASD subsample.

### Use of artificial intelligence tools

The authors used AI-based tools (ChatGPT and Grammarly) to assist with grammar and language editing of the manuscript. The authors reviewed and take full responsibility for the content of the manuscript. All scientific content and interpretations were determined by the authors.

## Results

### Descriptive statistics and scale score distributions

Across both samples, the ASD and non-ASD groups differed on the AQ-50 total score and all subscale scores, as indicated by Mann-Whitney *U* tests (all *p* < 0.001; see S1 Table for details). In line with expectations, participants in the ASD group endorsed higher levels of autistic traits than those in the non-ASD group. With respect to gender, we observed a pattern consistent with large-scale studies [23,49]: men tended to score higher on the AQ-50, and these gender differences were attenuated among autistic participants. Descriptive statistics are reported in Table 1, and score distributions by gender and diagnostic group are shown in Fig 1.

**Fig 1.**
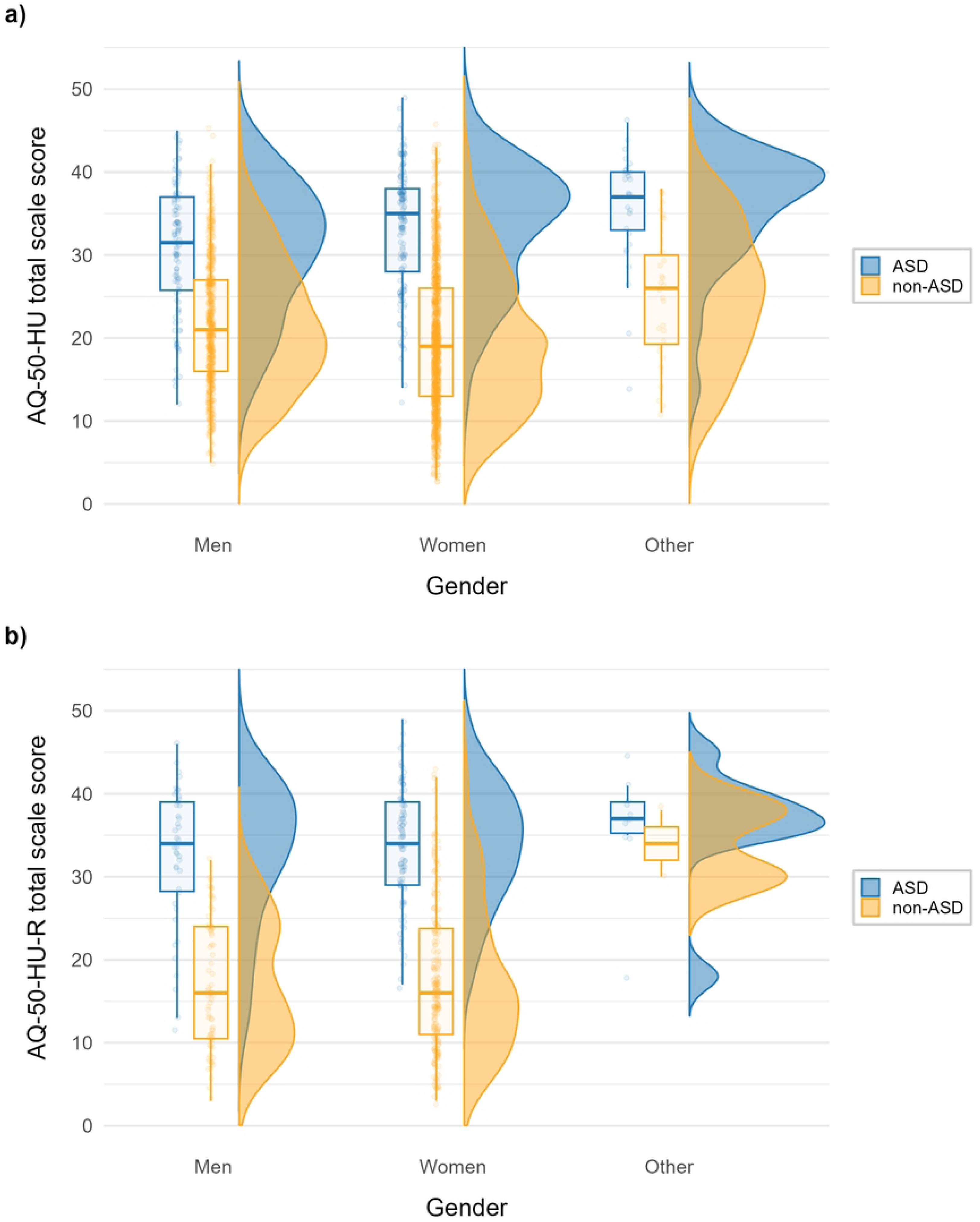
AQ-50 total score distributions across samples and diagnostic groups. (a) Sample 1 (*N* = 1967) assessed using the Hungarian version of the Autism Spectrum Quotient (AQ-50-HU), and (b) Sample 2 (*N* = 423), assessed using the revised Hungarian translation of the Autism Spectrum Quotient (AQ-50-HU-R).

### Internal consistency

Using the original Hungarian translation of the AQ-50 in Sample 1, both Cronbach’s alpha and McDonald’s omega exceeded conventional acceptability thresholds for the *social skill, communication,* and *attention switching* subscales, as well as for the total AQ-50-HU score. In contrast, the *attention to detail* and *imagination* subscales showed lower reliability estimates, falling below or near the recommended thresholds.

For the revised translation (AQ-50-HU-R) in Sample 2, reliability was higher, with all coefficients exceeding the recommended thresholds for both the total score and all subscales except *attention to detail.* Detailed reliability coefficients are presented in Table 2.

**Table 2.** Internal consistency and test-retest reliability of AQ-50 subscales and total score in Sample 1 and Sample 2.

|  | AQ-50-HU (Sample 1) |  |  | AQ-50-HU-R (Sample 2) |  |  |
| --- | --- | --- | --- | --- | --- | --- |
|  | Internal consistency<br>(*N* = 1967) |  | Test-retest<br>reliability<br>(*N* = 325) | Internal consistency<br>(*N* = 423) |  | Test-retest<br>reliability<br>(*N* = 97) |
| | Cronbach's<br>$\alpha$<br>[95% CI] | McDonald's<br>$\omega$<br>[95% CI] | Spearman's<br>correlation<br>($r_s$) | Cronbach's<br>$\alpha$<br>[95% CI] | McDonald's<br>$\omega$<br>[95% CI] | Spearman's<br>correlation<br>($r_s$) |
| **social skill** | 0.811<br>[0.799-0.823] | 0.814<br>[0.802-0.826] | 0.893*** | 0.851<br>[0.831-0.870] | 0.855<br>[0.835-0.875] | 0.908*** |
| **attention**<br>**switching** | 0.718<br>[0.700-0.737] | 0.722<br>[0.704-0.740] | 0.836*** | 0.824<br>[0.800-0.848] | 0.825<br>[0.801-0.850] | 0.851*** |
| **attention to**<br>**detail** | 0.583<br>[0.556-0.610] | 0.545<br>[0.519-0.571] | 0.812*** | 0.644<br>[0.595-0.694] | 0.646<br>[0.599-0.694] | 0.850*** |
| **communication** | 0.742<br>[0.724-0.759] | 0.735<br>[0.718-0.751] | 0.874*** | 0.844<br>[0.822-0.865] | 0.849<br>[0.828-0.870] | 0.932*** |
| **imagination** | 0.620<br>[0.594-0.646] | 0.623<br>[0.599-0.647] | 0.804*** | 0.716<br>[0.674-0.759] | 0.719<br>[0.679-0.758] | 0.911*** |
| **total scale score** | 0.886<br>[0.880-0.893] | 0.887<br>[0.880-0.893] | 0.935*** | 0.930<br>[0.922-0.938] | 0.932<br>[0.924-0.941] | 0.946*** |

AQ-50-HU = Hungarian version of the Autism Spectrum Quotient; AQ-50-HU-R = Revised translation of the Hungarian Autism Spectrum Quotient; CI = confidence interval.
\*\*\* $p < 0.001$ .

### Confirmatory factor analyses

#### Original translation (AQ-50-HU)

We first evaluated the original 5-factor model [20], which includes the *social skill, attention switching, attention to detail, communication,* and *imagination* subscales. The model did not demonstrate an acceptable fit to our data (*χ^2^*(1165) = 12665.833, *p* < 0.001; CFI = 0.898; TLI = 0.892; RMSEA = 0.071 (90% CI = [0.070; 0.072]); SRMR = 0.105). Next, we tested a unidimensional (1-factor) model, in which all items were specified to load on a single *general autistic traits* factor. This model also showed poor overall fit to the data (*χ^2^*(1175) = 14561.538, *p* < 0.001; CFI = 0.881; TLI = 0.876; RMSEA = 0.076 (90% CI = [0.075; 0.077]); SRMR = 0.107).

We additionally evaluated a bifactor structure for the AQ-50-HU, in which all items loaded on both a *general autistic traits* factor and their respective specific factors (*social skill, attention switching, attention to detail, communication,* and *imagination*). A schematic illustration of the bifactor model tested in the present study is shown in S1 Fig. This model showed acceptable overall fit to our data (*χ^2^*(1125) = 7514.523, *p* < 0.001; CFI = 0.943; TLI = 0.938; RMSEA = 0.054 (90% CI = [0.053; 0.055]); SRMR = 0.083). The overall reliability of the bifactor model was good (ω = 0.91). Regarding factor reliability, the *general autistic traits* factor demonstrated strong reliability (ω_H_ = 0.84), and the *attention to detail* specific factor showed acceptable reliability (ω_H_ = 0.55). The remaining specific factors exhibited weak reliability (ω_H_ = 0.003-0.29). The explained common variance (ECV = 0.61) indicated that 61% of the common variance was attributable to the *general autistic traits* factor. Construct replicability indices suggested excellent stability for the *general autistic traits* factor (H = 0.96), good stability for *attention to detail* (H = 0.83), *imagination* (H = 0.76), and *attention switching* (H = 0.70), while *communication* (H = 0.69) and *social skill* (H = 0.63) were below the recommended threshold.

Across all tested models, the bifactor solution estimated using binary scoring provided the best overall fit to the present data. Accordingly, we retained this model and examined whether the factor structure would replicate in an independent sample. In Sample 2, the bifactor model again demonstrated good fit (*χ^2^*(1125) = 1744.392, *p* < 0.001; CFI = 0.988; TLI = 0.987; RMSEA = 0.036 (90% CI = [0.033; 0.039]); SRMR = 0.085), with highly comparable indices of reliability (ω_H_) and construct replicability (H). A detailed summary of the CFA fit indices is presented in the Supporting Information (S2 and S3 Tables).

#### Revised translation (AQ-50-HU-R)

For the revised translation (AQ-50-HU-R), we retained the bifactor specification and used Sample 2 to evaluate whether the factor structure differed under the revised item set, in which the seven previously identified items were replaced with their updated translations. The bifactor model demonstrated acceptable fit to the data (*χ^2^*(1125) = 1650.433, *p* < 0.001; CFI = 0.991; TLI = 0.990; RMSEA = 0.033 (90% CI = [0.030; 0.037]); SRMR = 0.083). Overall model reliability was high (ω = 0.95). Standardized factor loadings varied substantially across items, ranging from |0.01| to |0.93|. One item (item 29) had a nonsignificant loading on the *general autistic traits* factor, and two items (items 30 and 49) showed negative general-factor loadings despite all items being coded in the same direction. Notably, all three items belonged to the *attention to detail* specific factor. Across the five specific factors, each included at least one item with either a nonsignificant or a negative loading. Standardized loadings and the proportion of total item variance explained are reported in Table 3.

**Table 3.** Standardized factor loadings of the revised translation of the Hungarian Autism Spectrum Quotient (AQ-50-HU-R) based on Sample 2 (*N* = 423).

| Item | Factor loadings | | | | | | Explained variance ($R^2$) |
| --- | --- | --- | --- | --- | --- | --- | --- |
|  | *social skill* | *attention switching* | *attention to detail* | *communication* | *imagination* | *general autistic traits* |  |
| AQ-1 | 0.51 |  |  |  |  | 0.61 | 0.63 |
| AQ-11 | 0.17 |  |  |  |  | 0.91 | 0.86 |
| AQ-13 | 0.28 |  |  |  |  | 0.45 | 0.28 |
| AQ-15 | 0.13 |  |  |  |  | 0.67 | 0.47 |
| AQ-22 | 0.26 |  |  |  |  | 0.80 | 0.71 |
| AQ-36 | -0.19 |  |  |  |  | 0.86 | 0.77 |
| AQ-44 | 0.58 |  |  |  |  | 0.77 | 0.92 |
| AQ-45 | -0.22 |  |  |  |  | 0.91 | 0.88 |
| AQ-47 | 0.54 |  |  |  |  | 0.66 | 0.72 |
| AQ-48 | **-0.05** |  |  |  |  | 0.55 | 0.30 |
| AQ-2 |  | 0.43 |  |  |  | 0.59 | 0.53 |
| AQ-4 |  | **-0.01** |  |  |  | 0.71 | 0.51 |
| AQ-10 |  | 0.14 |  |  |  | 0.74 | 0.57 |
| AQ-16 |  | 0.17 |  |  |  | 0.69 | 0.51 |
| AQ-25 |  | 0.58 |  |  |  | 0.62 | 0.71 |
| AQ-32 |  | 0.30 |  |  |  | 0.54 | 0.38 |
| AQ-34 |  | 0.63 |  |  |  | 0.55 | 0.70 |
| AQ-37 |  | 0.23 |  |  |  | 0.78 | 0.66 |
| AQ-43 |  | 0.55 |  |  |  | 0.36 | 0.43 |
| AQ-46 |  | 0.16 |  |  |  | 0.76 | 0.61 |
| AQ-5 |  |  | 0.31 |  |  | 0.60 | 0.46 |
| AQ-6 |  |  | 0.52 |  |  | 0.51 | 0.54 |
| AQ-9 |  |  | 0.68 |  |  | 0.42 | 0.64 |
| AQ-12 |  |  | 0.44 |  |  | 0.49 | 0.43 |
| AQ-19 |  |  | 0.62 |  |  | 0.39 | 0.54 |
| AQ-23 |  |  | 0.35 |  |  | 0.39 | 0.28 |
| AQ-28 |  |  | **0.04** |  |  | 0.71 | 0.51 |
| AQ-29 |  |  | 0.57 |  |  | **-0.02** | 0.33 |
| AQ-30 |  |  | 0.42 |  |  | -0.30 | 0.26 |
| AQ-49 |  |  | 0.56 |  |  | -0.14 | 0.34 |
| AQ-7 |  |  |  | 0.34 |  | 0.73 | 0.65 |
| AQ-17 |  |  |  | -0.52 |  | 0.67 | 0.72 |

| Item | Factor loadings |  |  |  |  |  | Explained variance (R<sup>2</sup>) |
| --- | --- | --- | --- | --- | --- | --- | --- |
|  | *social skill* | *attention switching* | *attention to detail* | *communication* | *imagination* | *general autistic traits* |  |
| AQ-18 |  |  |  | 0.63 |  | 0.34 | 0.51 |
| AQ-26 |  |  |  | -0.18 |  | 0.87 | 0.79 |
| AQ-27 |  |  |  | **0.10** |  | 0.83 | 0.70 |
| AQ-31 |  |  |  | 0.17 |  | 0.80 | 0.68 |
| AQ-33 |  |  |  | 0.21 |  | 0.75 | 0.60 |
| AQ-35 |  |  |  | 0.26 |  | 0.67 | 0.52 |
| AQ-38 |  |  |  | -0.24 |  | 0.93 | 0.92 |
| AQ-39 |  |  |  | 0.37 |  | 0.86 | 0.87 |
| AQ-3 |  |  |  |  | 0.81 | 0.22 | 0.70 |
| AQ-8 |  |  |  |  | 0.75 | 0.31 | 0.65 |
| AQ-14 |  |  |  |  | 0.60 | 0.30 | 0.45 |
| AQ-20 |  |  |  |  | 0.25 | 0.70 | 0.55 |
| AQ-21 |  |  |  |  | 0.22 | 0.42 | 0.23 |
| AQ-24 |  |  |  |  | **0.01** | 0.41 | 0.17 |
| AQ-40 |  |  |  |  | 0.19 | 0.54 | 0.33 |
| AQ-41 |  |  |  |  | **-0.08** | 0.68 | 0.47 |
| AQ-42 |  |  |  |  | 0.30 | 0.66 | 0.52 |
| AQ-50 |  |  |  |  | 0.35 | 0.64 | 0.53 |

AQ = Autism Spectrum Quotient; R<sup>2</sup> = explained variance; bold values indicate
nonsignificant loadings ( $p \geq 0.05$ ).

Consistent with the bifactor findings described above, the *general autistic traits* factor exhibited strong hierarchical reliability (ω_H_ = 0.90). In contrast, the domain-specific factors showed weak hierarchical reliability (ω_H_ = 0.02-0.49), suggesting limited common variance beyond the general factor. Construct replicability indices were consistent with this pattern, indicating excellent stability for the *general autistic traits* factor (H = 0.98), and good stability for *imagination* (H = 0.80) and *attention to detail* (H = 0.78), but weaker replicability for *communication* (H = 0.61), *attention switching* (H = 0.67), and *social skill* (H = 0.61). Explained common variance (ECV = 0.71) further supported the predominance of the general factor, indicating that 71% of the common variance was attributable to the *general autistic traits* dimension. Detailed model fit and reliability indices are reported in Table 4.

**Table 4.**
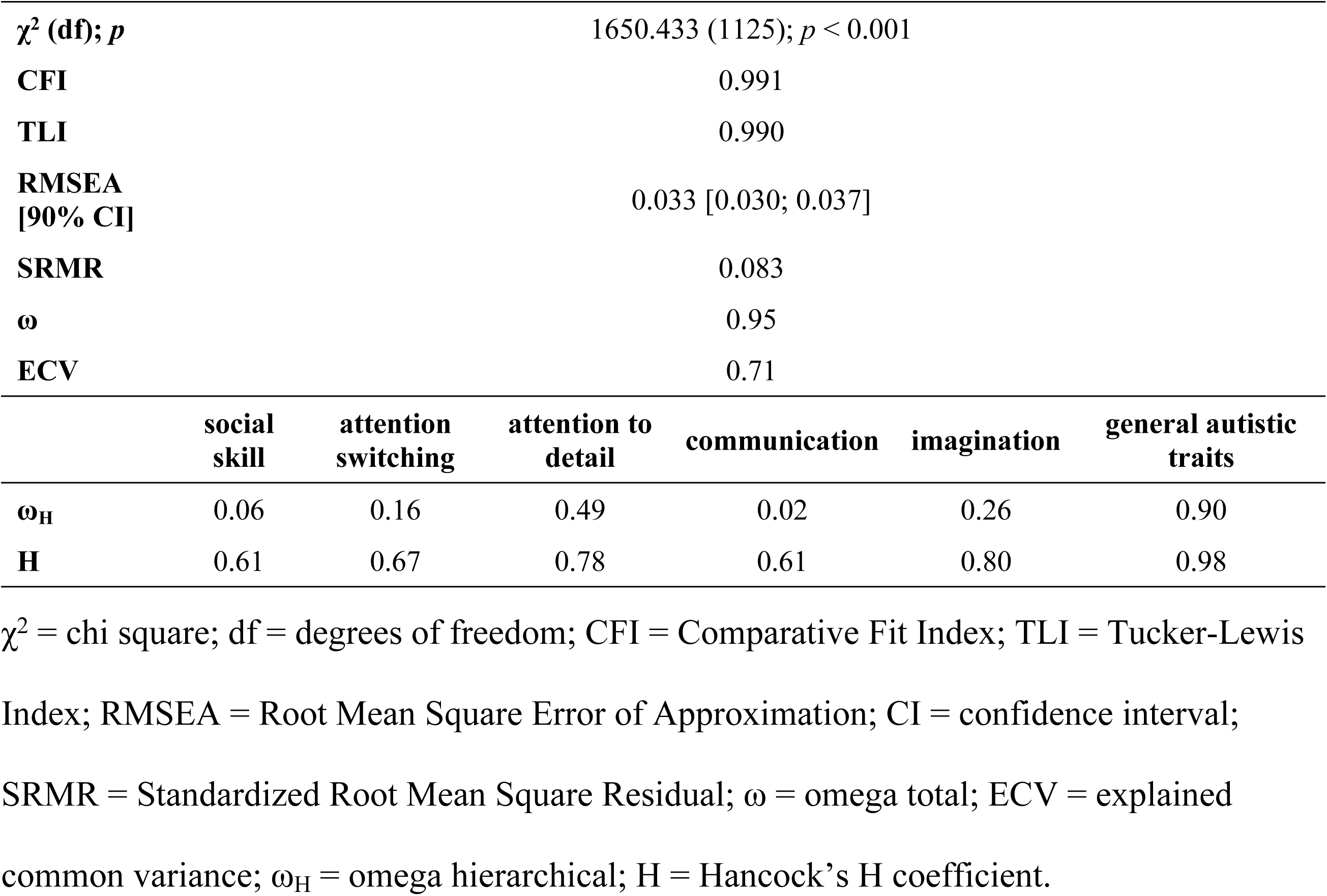
Bifactor model of the revised translation of the Hungarian Autism Spectrum Quotient (AQ-50-HU-R) based on Sample 2 (*N* = 423).

| $\chi^2$ (df); $p$ | 1650.433 (1125); $p < 0.001$ | | | | | |
| --- | --- | --- | --- | --- | --- | --- |
| CFI | 0.991 |  |  |  |  |  |
| TLI | 0.990 |  |  |  |  |  |
| RMSEA<br>[90% CI] | 0.033 [0.030; 0.037] |  |  |  |  |  |
| SRMR | 0.083 |  |  |  |  |  |
| $\omega$ | 0.95 | | | | | |
| ECV | 0.71 |  |  |  |  |  |
|  | social<br>skill | attention<br>switching | attention to<br>detail | communication | imagination | general autistic<br>traits |
| $\omega_H$ | 0.06 | 0.16 | 0.49 | 0.02 | 0.26 | 0.90 |
| H | 0.61 | 0.67 | 0.78 | 0.61 | 0.80 | 0.98 |

$\chi^2$ = chi square; df = degrees of freedom; CFI = Comparative Fit Index; TLI = Tucker-Lewis Index; RMSEA = Root Mean Square Error of Approximation; CI = confidence interval; SRMR = Standardized Root Mean Square Residual; $\omega$ = omega total; ECV = explained common variance; $\omega_H$ = omega hierarchical; H = Hancock's H coefficient.

### Test-retest reliability

To evaluate the test-retest reliability and temporal stability of the AQ-50-HU and AQ-50-HU-R, participants completed the questionnaire again after 6 months in Sample 1 and 1 month in Sample 2. Given the ordinal categorical nature of the data, we used Spearman’s rank correlations (*r_s_*) to examine associations between scores obtained at the two time points. For both the AQ-50-HU and AQ-50-HU-R, the total score and all subscale scores showed strong, statistically significant test-retest correlations, indicating excellent temporal stability. Detailed coefficients are reported in Table 2.

### Screening accuracy of the AQ-50 total scores

To evaluate the predictive value and clinical utility of the AQ-50 total scores in distinguishing between ASD and non-ASD participants, we conducted receiver operating characteristic (ROC) analyses. In both samples, participants with unverified ASD status were initially excluded. Based on the Youden index, the optimal cutoff for discriminating between ASD and non-ASD participants was 28 in Sample 1 and 25 in Sample 2. These thresholds yielded areas under the curve (AUC) of 0.880 and 0.906, respectively, indicating good discriminative accuracy and supporting the utility of the AQ-50 as a screening instrument. Detailed performance indices are reported in Table 5 and illustrated in Fig 2.

**Fig 2.**
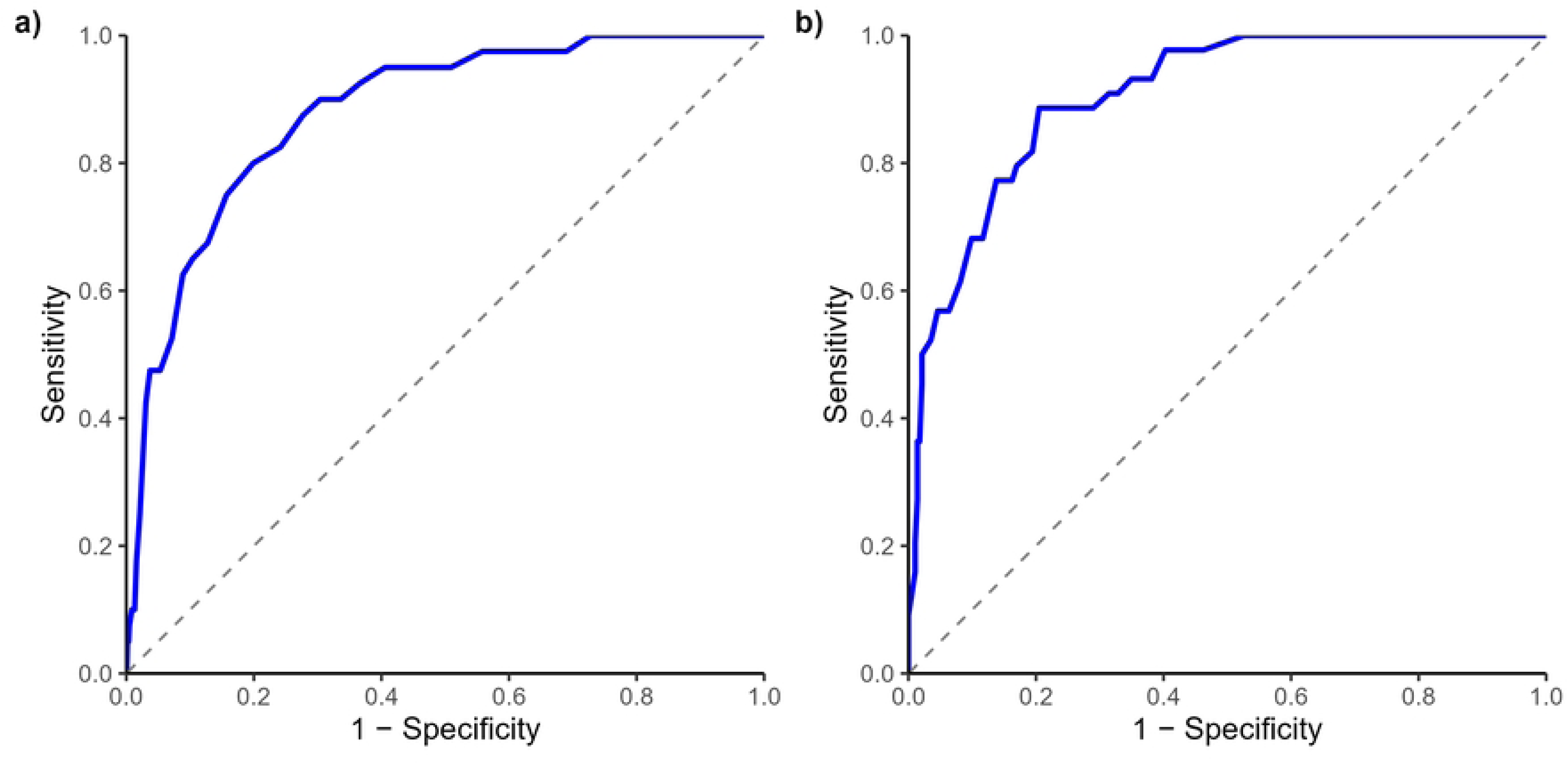
Receiver operating characteristic curves for AQ-50 total scores. Receiver operating characteristic (ROC) curves for AQ-50 total scores distinguishing clinically verified ASD participants from non-ASD participants. (a) AQ-50-HU total score in Sample 1 (*N* = 1733; AUC = 0.880, 95% CI = [0.831, 0.929]). (b) AQ-50-HU-R total score in Sample 2 (*N* = 327; AUC = 0.906, 95% CI = [0.865, 0.947]).

**Table 5.** Accuracy of AQ-50 total scores in discriminating clinically verified ASD cases from non-ASD cases in Sample 1 (*N* = 1733) and Sample 2 (*N* = 327).

|  | **AQ-50-HU**<br>**($N = 1733$)** | **AQ-50-HU-R**<br>**($N = 327$)** |
| --- | --- | --- |
| **Youden-index optimal cut-off** | **28** | **25** |
| **AUC** | 0.880 | 0.906 |
| **[95% CI]** | [0.831-0.929] | [0.865-0.947] |
| **Sensitivity** | 0.800 | 0.886 |
| **Specificity** | 0.801 | 0.795 |
| PPV | 0.087 | 0.402 |
| NPV | 0.994 | 0.978 |
| ACC | 0.801 | 0.807 |

AQ-50-HU = Hungarian version of the Autism Spectrum Quotient; AQ-50-HU-R = Revised translation of the Hungarian Autism Spectrum Quotient; AUC = area under the receiver operating characteristic curve; CI = confidence interval; PPV = positive predictive value; NPV = negative predictive value; ACC = accuracy.

We then repeated the ROC analyses using a broader group definition, in which we jointly classified participants with documented or self-reported ASD diagnoses in the ASD group, and participants reporting no autism diagnosis as the comparison group. Under this definition, the optimal cutoffs remained unchanged, yielding AUCs of 0.854 and 0.907 for Samples 1 and 2, respectively. Detailed performance indices are reported in S4 Table.

### Convergent validity

We examined convergent validity by assessing associations between AQ-50 scores and the ADOS A (*language and communication*), B (*reciprocal social interaction*), and combined (A+B) domain scores, using Spearman’s correlations (*r_s_*). Among all tested associations, only the AQ-50 *social skill* subscale showed a weak, significant positive correlation with the ADOS-B domain. All remaining correlations were nonsignificant (see Table 6), suggesting limited convergence between self-reported autistic traits and clinician-rated features.

**Table 6.**
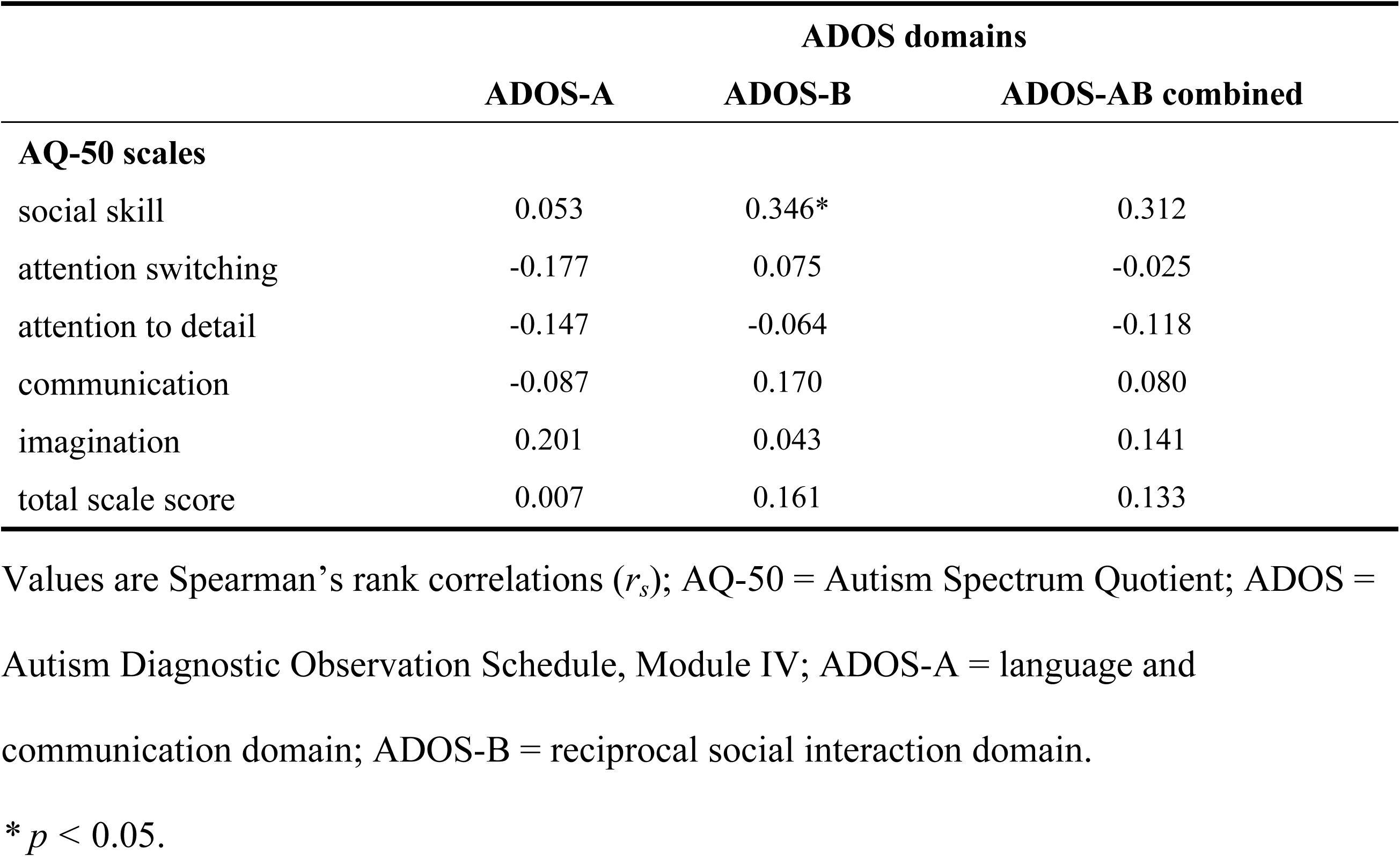
Associations between AQ-50 scores and ADOS scores in the clinical ASD subsample (*N* = 40) from Sample 1.

|  | ADOS domains |  |  |
| --- | --- | --- | --- |
|  | ADOS-A | ADOS-B | ADOS-AB combined |
| **AQ-50 scales** |  |  |  |
| social skill | 0.053 | 0.346* | 0.312 |
| attention switching | -0.177 | 0.075 | -0.025 |
| attention to detail | -0.147 | -0.064 | -0.118 |
| communication | -0.087 | 0.170 | 0.080 |
| imagination | 0.201 | 0.043 | 0.141 |
| total scale score | 0.007 | 0.161 | 0.133 |

Values are Spearman's rank correlations ( $r_s$ ); AQ-50 = Autism Spectrum Quotient; ADOS = Autism Diagnostic Observation Schedule, Module IV; ADOS-A = language and communication domain; ADOS-B = reciprocal social interaction domain.
\* $p < 0.05$ .

## Discussion

The present study examined the psychometric properties, factorial structure, and clinical applicability of the revised Hungarian version of the *Autism Spectrum Quotient* (AQ-50-HU-R) in adult samples with and without an autism diagnosis. Overall, the findings suggest that the AQ-50-HU-R can serve as a reliable measure of self-reported autistic traits and a clinically informative screening instrument when interpreted at the total score level. At the same time, our results do not support confidently interpreting the original five subscales as robust, psychometrically reliable dimensions.

The distributional characteristics of our samples were comparable with those reported in large-scale international studies [23,49]. Men tended to score higher on the AQ-50 than women, and these differences were attenuated among autistic participants. Baron-Cohen [50] interpreted this pattern within the framework of the extreme male brain (EMB) theory of autism, according to which neurotypical males tend to show stronger systemizing, whereas females tend to show stronger empathizing. Within this framework, autism is conceptualized as a shift toward a more systemizing cognitive style, which may attenuate typical sex differences. However, an alternative interpretation is that the AQ-50 may better capture a more traditionally male autism phenotype, may align more closely with stereotypically male trait expressions, or may be less sensitive to camouflaging behaviors, which are often discussed in relation to the female autism phenotype [4,10]. Consistent with this possibility, evidence suggests that AQ-50 items are not fully invariant across sex or gender and may operate differently for different respondents. Specifically, women may be more likely to endorse items related to social and communication skills [51].

With regard to factor structure, neither the original five-factor model [20] nor a unidimensional model was supported, as both showed suboptimal fit. By contrast, a bifactor solution provided the best overall representation of the data. This model comprised a *general autistic traits* factor and five domain-specific factors (*social skill, attention switching, attention to detail, communication,* and *imagination*). At the same time, it is important to emphasize that, across all tested bifactor models, the *general autistic traits* factor accounted for most of the shared variance among items. *Attention to detail* retained some residual variance, whereas the remaining domain-specific factors contributed little to no interpretable variance above the general dimension. Taken together, these findings suggest that the AQ-50 functions primarily as a measure of broad autistic traits rather than as a profile instrument capturing five trait domains. This pattern is consistent with previous research showing that the original five-factor structure often does not replicate and that the AQ-50 factor structure varies considerably across languages, scoring formats, and sample types [24,26,52]. In our final model, items 29, 30, and 49, all belonging to the *attention to detail* domain, appeared to perform poorly as indicators of autistic traits and may require revision. Similar difficulties with these items have also been reported in other language versions of the AQ-50 [53,54]. Our bifactor solution closely resembles that reported by Murray et al. [29], who likewise concluded that AQ-50 items primarily reflect a general autistic trait dimension rather than separable domain-specific constructs.

Consistent with previous international studies [27,35,55,56], the total AQ-50 score demonstrated good internal consistency in both the original and revised Hungarian translations and across both samples. However, we did not observe this pattern for the attention to detail and imagination subscales, which showed more variable reliability, ranging from acceptable to below the accepted threshold. These findings further support the interpretation of the total score as a reliable indicator of self-reported autistic traits. Regarding test-retest reliability, both the total score and the subscale scores showed strong temporal stability over intervals of 1 and 6 months. This finding is conceptually consistent with longitudinal evidence that autistic traits show relative temporal stability and may represent developmentally stable dispositional characteristics [55,57].

According to our findings, self-reported autistic traits and clinician-rated behavioral features of autism were essentially uncorrelated. Although this may appear counterintuitive at first glance, it is consistent with previous studies reporting similarly weak associations between AQ-50 scores and observer-based autism assessments [32,58,59]. One possible explanation is that the AQ-50 may be more sensitive to the subjective experience of difficulties and camouflaging, whereas the ADOS is designed to capture autistic characteristics as evident to observers [60]. Although the diagnosis of autism requires a comprehensive clinical assessment based on developmental history, clinical interviews, and behavioral observations, the AQ-50, as a self-report questionnaire, serves as a valuable tool for understanding the subjective experiences, quality of life, and needs of autistic individuals.

In terms of clinical utility, analyses of clinically verified ASD subsamples indicated that the total AQ-50 score demonstrated good discriminative accuracy in distinguishing ASD from non-ASD participants. Specifically, the empirically derived cutoff score of 25 provided the best balance between sensitivity and specificity for the AQ-50-HU-R total score in our Sample 2. This threshold is substantially lower than the original cutoff of 32 [20] and is consistent with several international studies reporting that a cutoff around 25 may be optimal for discriminating between autistic and non-autistic individuals [30–33,58,61]. At the same time, the combination of high negative predictive values and lower positive predictive values indicates that a substantial proportion of individuals scoring above the cutoff would not necessarily meet diagnostic criteria for ASD. Importantly, PPV and NPV should not be interpreted as stable properties of the questionnaire, as both indices are strongly influenced by prevalence and sampling design. Taken together, these findings suggest that the AQ-50-HU-R may be more useful for identifying individuals who are likely to require further diagnostic evaluation than for confirming ASD. This interpretation is consistent with its intended role as a screening instrument and further underscores that it should not be used as a stand-alone diagnostic tool. It is also important to note that phenotypically similar conditions, including characteristics of ADHD, anxiety disorders, OCD, schizophrenia-spectrum conditions, eating disorders, and personality disorders, may also be associated with elevated AQ-50 scores, suggesting that autistic characteristics are not specific to the autism spectrum [32,58,62–64].

### Limitations and further directions

Several limitations of our study should be acknowledged. First, the reliance on predominantly online convenience samples limits the generalizability of the findings to the broader Hungarian adult population. Second, only a small proportion of self-reported ASD diagnoses could be clinically verified (*N_1_* = 40; *N_2_*= 44), and ADOS scores were available only for Sample 1. Because the present study primarily examined the psychometric properties and factorial structure of the Hungarian AQ-50, certain aspects of its broader validation remain unresolved. Future research should place greater emphasis on convergent and discriminant validity. Convergent validity could be examined by comparing the AQ-50 with other self-report measures of autistic traits and social differences, such as the SRS-2 [19] and the RAADS-R [18]. Discriminant validity, in turn, could be assessed by examining associations with related but distinct constructs, such as social anxiety. In addition, the present study did not examine measurement invariance. Future research should therefore test whether the Hungarian AQ-50 functions equivalently across gender groups and across clinical and non-clinical samples.

## Conclusions

In conclusion, the present findings suggest that the AQ-50-HU-R may serve as a reliable instrument for assessing variation in autistic traits among Hungarian-speaking adults and for screening individuals who may benefit from a more comprehensive autism evaluation. For screening purposes, we recommend a total score cutoff of 25. The original subscale scores should be interpreted with caution and are not recommended for individual-level clinical decision-making, whereas the bifactor model supports interpreting the total score as a measure of self-reported autistic traits. We recommend the questionnaire not be used as a stand-alone diagnostic tool, as elevated scores may also occur in other neurodevelopmental or psychiatric conditions. Positive screening results should therefore be followed by a comprehensive clinical assessment, including developmental history, clinical interview, and standardized observational measures, in accordance with current clinical guidelines.

## Data Availability

The minimal data set and statistical code supporting the CFA and ROC analyses reported in this study are available on the Open Science Framework (OSF) at the following anonymous view-only link: https://osf.io/6keb5/overview?view_only=2b24e498a3d94801a1eb2c3ed8a9fb3d

## Acknowledgments

None.

## Supporting information

S1 Fig. Schematic illustration of the AQ-50 bifactor model.

Each of the AQ-50 items loads on the *general autistic traits* factor and on one specific factor.

S1 Table. Shapiro-Wilk normality tests and Mann-Whitney U tests for demographic variables and AQ-50 scale scores.

*W* = Shapiro-Wilk test statistic; *U* = Mann-Whitney U statistic; ASD = autism spectrum disorder; AQ-50-HU = Hungarian version of the Autism Spectrum Quotient; AQ-50-HU-R = Revised translation of the Hungarian Autism Spectrum Quotient; Significant Shapiro-Wilk tests indicate deviation from normality. *** *p* < 0.001; ** *p* < 0.01.

S2 Table. Fit indices of the tested factor models of the AQ-50-HU in Sample 1.

AQ-50-HU = Hungarian version of the Autism Spectrum Quotient; χ^2^ = chi square; df = degrees of freedom; CFI = Comparative Fit Index; TLI = Tucker-Lewis Index; RMSEA = Root Mean Square Error of Approximation; CI = confidence interval; SRMR = Standardized Root Mean Square Residual; ω = omega total; ECV = explained common variance; ω_H_ = omega hierarchical; H = Hancock’s H coefficient.

S3 Table. Fit indices of the bifactor model of the original Hungarian translation (AQ-50-HU) in Sample 2.

AQ-50-HU = Hungarian version of the Autism Spectrum Quotient; χ^2^ = chi square; df = degrees of freedom; CFI = Comparative Fit Index; TLI = Tucker-Lewis Index; RMSEA = Root Mean Square Error of Approximation; CI = confidence interval; SRMR = Standardized Root Mean Square Residual; ω = omega total; ECV = explained common variance; ω_H_ = omega hierarchical; H = Hancock’s H coefficient.

S4 Table. Accuracy of AQ-50 total scores in discriminating self-reported ASD cases from non-ASD cases.

AQ-50-HU = Hungarian version of the Autism Spectrum Quotient; AQ-50-HU-R = Revised translation of the Hungarian Autism Spectrum Quotient; AUC = area under the receiver operating characteristic curve; CI = confidence interval; PPV = positive predictive value; NPV = negative predictive value; ACC = accuracy.

## Notes

### Competing Interest Statement

The authors have declared no competing interest.

### Author Declarations

Ethical approval was granted by the Semmelweis University Regional and Institutional Committee of Science and Research Ethics and the Medical Research Council (approval numbers: RKEB 158/2021 RKEB 159/2021 and BM/14269-3/2025). Before data collection, all participants provided electronic written informed consent.

## References

1. American Psychiatric Association. Diagnostic and Statistical Manual of Mental Disorders: DSM-5-TR. 5th ed, text rev. Washington, DC: Author; 2022. Available: 10.1176/appi.books.9780890425787

2. Lever AG, Geurts HM. Psychiatric Co-occurring Symptoms and Disorders in Young, Middle-Aged, and Older Adults with Autism Spectrum Disorder. J Autism Dev Disord. 2016;46: 1916–1930. doi:10.1007/s10803-016-2722-8

3. Mutluer T, Aslan Genç H, Özcan Morey A, Yapici Eser H, Ertinmaz B, Can M, et al. Population-Based Psychiatric Comorbidity in Children and Adolescents With Autism Spectrum Disorder: A Meta-Analysis. Front Psychiatry. 2022;13: 856208. doi:10.3389/fpsyt.2022.856208

4. Lai M-C, Lombardo MV, Baron-Cohen S. Autism. The Lancet. 2014;383: 896–910. doi:10.1016/S0140-6736(13)61539-1

5. Maenner MJ, Warren Z, Williams AR, Amoakohene E, Bakian AV, Bilder DA, et al. Prevalence and Characteristics of Autism Spectrum Disorder Among Children Aged 8 Years - Autism and Developmental Disabilities Monitoring Network, 11 Sites, United States, 2020. Morb Mortal Wkly Rep Surveill Summ. 2023;72: 1–14. doi:10.15585/mmwr.ss7202a1

6. Shaw KA, Williams S, Patrick ME, Valencia-Prado M, Durkin MS, Howerton EM, et al. Prevalence and Early Identification of Autism Spectrum Disorder Among Children Aged 4 and 8 Years - Autism and Developmental Disabilities Monitoring Network, 16 Sites, United States, 2022. Morb Mortal Wkly Rep Surveill Summ. 2025;74: 1–22. doi:10.15585/mmwr.ss7402a1

7. Brugha T, Tyrer F, Leaver A, Lewis S, Seaton S, Morgan Z, et al. Testing adults by questionnaire for social and communication disorders, including autism spectrum disorders, in an adult mental health service population. Int J Methods Psychiatr Res. 2020;29: e1814. doi:10.1002/mpr.1814

8. Nyrenius J, Eberhard J, Ghaziuddin M, Gillberg C, Billstedt E. Prevalence of Autism Spectrum Disorders in Adult Outpatient Psychiatry. J Autism Dev Disord. 2022;52: 3769–3779. doi:10.1007/s10803-021-05411-z

9. O’Nions E, Petersen I, Buckman JEJ, Charlton R, Cooper C, Corbett A, et al. Autism in England: assessing underdiagnosis in a population-based cohort study of prospectively collected primary care data. Lancet Reg Health - Eur. 2023;29: 100626. doi:10.1016/j.lanepe.2023.100626

10. Hull L, Petrides KV, Mandy W. The Female Autism Phenotype and Camouflaging: a Narrative Review. Rev J Autism Dev Disord. 2020;7: 306–317. doi:10.1007/s40489-020-00197-9

11. Bishop-Fitzpatrick L, Minshew NJ, Eack SM. A Systematic Review of Psychosocial Interventions for Adults with Autism Spectrum Disorders. J Autism Dev Disord. 2013;43: 687–694. doi:10.1007/s10803-012-1615-8

12. MacKenzie KT, Theodat A, Beck KB, Conner CM, Mazefsky CA, Eack SM. Correlates of quality of life in autistic individuals. Res Autism Spectr Disord. 2024;115: 102401. doi:10.1016/j.rasd.2024.102401

13. Farkas K, Bálint M, Baráth K, Gallai M, Lakos E, Lisincki A, et al. Az autizmus diagnosztikája és ellátása felnőttkorban. Psychiatr Hung. 2024;39: 258–285.

14. National Institute for Health and Care Excellence. NICE Guideline. Autism spectrum disorder in adults: diagnosis and management. 2021. Available: https://www.nice.org.uk/guidance/cg142

15. Lord C, Rutter M, Goode S, Heemsbergen J, Jordan H, Mawhood L, et al. Autism Diagnostic Observation Schedule: A standardized observation of communicative and social behavior. J Autism Dev Disord. 1989;19: 185–212. doi:10.1007/BF02211841

16. Lord C, Rutter M, DiLavore PS, Risi S. Autism Diagnostic Observation Schedule (ADOS). Los Angeles, CA: Western Psychological Services; 2000.

17. Le Couteur A, Lord C, Rutter M. The Autism Diagnostic Interview-Revised. Los Angeles: Western Psychological Services; 2003.

18. Ritvo RA, Ritvo ER, Guthrie D, Ritvo MJ, Hufnagel DH, McMahon W, et al. The Ritvo Autism Asperger Diagnostic Scale-Revised (RAADS-R): A Scale to Assist the Diagnosis of Autism Spectrum Disorder in Adults: An International Validation Study. J Autism Dev Disord. 2011;41: 1076–1089. doi:10.1007/s10803-010-1133-5

19. Constantino JN, Gruber CP. Social Responsiveness Scale, Second Edition (SRS-2). Torrance: Western Psychological Services; 2012.

20. Baron-Cohen S, Wheelwright S, Skinner R, Martin J, Clubley E. The Autism-Spectrum Quotient (AQ): Evidence from Asperger Syndrome/High-Functioning Autism, Males and Females, Scientists and Mathematicians. J Autism Dev Disord. 2001;31: 5–17.

21. Happé F, Frith U. Annual Research Review: Looking back to look forward – changes in the concept of autism and implications for future research. J Child Psychol Psychiatry. 2020;61: 218–232. doi:10.1111/jcpp.13176

22. Wainer AL, Ingersoll BR, Hopwood CJ. The Structure and Nature of the Broader Autism Phenotype in a Non-clinical Sample. J Psychopathol Behav Assess. 2011;33: 459–469. doi:10.1007/s10862-011-9259-0

23. Ruzich E, Allison C, Smith P, Watson P, Auyeung B, Ring H, et al. Measuring autistic traits in the general population: a systematic review of the Autism-Spectrum Quotient (AQ) in a nonclinical population sample of 6,900 typical adult males and females. Mol Autism. 2015;6: 2. doi:10.1186/2040-2392-6-2

24. English MCW, Gignac GE, Visser TAW, Whitehouse AJO, Maybery MT. A comprehensive psychometric analysis of autism-spectrum quotient factor models using two large samples: Model recommendations and the influence of divergent traits on total-scale scores. Autism Res. 2020;13: 45–60. doi:10.1002/aur.2198

25. Zain E, Fukui N, Watanabe Y, Hashijiri K, Motegi T, Ogawa M, et al. The three-factor structure of the Autism-Spectrum Quotient Japanese version in pregnant women. Front Psychiatry. 2023;14: 1275043. doi:10.3389/fpsyt.2023.1275043

26. Zhu Y, Mu W, Chirica MG, Berenbaum H. Testing a theory-driven factor structure of the autism-spectrum quotient. Autism Res. 2022;15: 1710–1718. doi:10.1002/aur.2763

27. Dolfi A, Faur D, Scălcău M-R, Sorescu E-M, Ciumăgeanu M-D, Tudose C. Validation of the Romanian Version of Autism Spectrum Quotient (AQ) and Empathy Quotient (EQ) in the General Population. Psychiatr Q. 2025;96: 803–818. doi:10.1007/s11126-025-10144-8

28. Moshirian Farahi SMM, Leth-Steensen C. Latent profile analysis of autism spectrum quotient. Curr Psychol. 2023;42: 30029–30036. doi:10.1007/s12144-022-03990-3

29. Murray AL, McKenzie K, Kuenssberg R, Booth T. Do the Autism Spectrum Quotient (AQ) and Autism Spectrum Quotient Short Form (AQ-S) Primarily Reflect General ASD Traits or Specific ASD Traits? A Bi-Factor Analysis. Assessment. 2017;24: 444–457. doi:10.1177/1073191115611230

30. Woodbury-Smith MR, Robinson J, Wheelwright S, Baron-Cohen S. Screening Adults for Asperger Syndrome Using the AQ: A Preliminary Study of its Diagnostic Validity in Clinical Practice. J Autism Dev Disord. 2005;35: 331–335. doi:10.1007/s10803-005-3300-7

31. Yoshinaga K, Egawa J, Watanabe Y, Kasahara H, Sugimoto A, Someya T. Usefulness of the autism spectrum quotient (AQ) in screening for autism spectrum disorder and social communication disorder. BMC Psychiatry. 2023;23: 831. doi:10.1186/s12888-023-05362-y

32. Ashwood KL, Gillan N, Horder J, Hayward H, Woodhouse E, McEwen FS, et al. Predicting the diagnosis of autism in adults using the Autism-Spectrum Quotient (AQ) questionnaire. Psychol Med. 2016;46: 2595–2604. doi:10.1017/S0033291716001082

33. Bezemer ML, Blijd-Hoogewys EMA, Meek-Heekelaar M. The Predictive Value of the AQ and the SRS-A in the Diagnosis of ASD in Adults in Clinical Practice. J Autism Dev Disord. 2021;51: 2402–2415. doi:10.1007/s10803-020-04699-7

34. Arslan RC, Walther MP, Tata CS. formr: A study framework allowing for automated feedback generation and complex longitudinal experience-sampling studies using R. Behav Res Methods. 2020;52: 376–387. doi:10.3758/s13428-019-01236-y

35. Stevenson JL, Hart KR. Psychometric Properties of the Autism-Spectrum Quotient for Assessing Low and High Levels of Autistic Traits in College Students. J Autism Dev Disord. 2017;47: 1838–1853. doi:10.1007/s10803-017-3109-1

36. Austin EJ. Personality correlates of the broader autism phenotype as assessed by the Autism Spectrum Quotient (AQ). Personal Individ Differ. 2005;38: 451–460. doi:10.1016/j.paid.2004.04.022

37. JASP Team. JASP. 2025. Available: https://jasp-stats.org/

38. R Core Team. R: A language and environment for statistical computing. Vienna, Austria: R Foundation for Statistical Computing; 2024. Available: https://www.R-project.org/

39. Posit team. RStudio: Integrated Development Environment for R. Boston, MA: Posit Software, PBC; 2025. Available: http://www.posit.co/

40. Wickham H, François R, Henry L, Müller K, Vaughan D. dplyr: A Grammar of Data Manipulation. 2023. Available: https://CRAN.R-project.org/package=dplyr

41. Dueber D. BifactorIndicesCalculator: Bifactor Indices Calculator. 2021. Available: https://CRAN.R-project.org/package=BifactorIndicesCalculator

42. Tiedemann F. gghalves: Compose Half-Half Plots Using Your Favourite Geoms. 2026. Available: https://github.com/erocoar/gghalves

43. Wickham H. ggplot2: Elegant Graphics for Data Analysis. 2nd ed. 2016. Cham: Springer International Publishing : Imprint: Springer; 2016. doi:10.1007/978-3-319-24277-4

44. Rosseel Y. **lavaan** : An *R* Package for Structural Equation Modeling. J Stat Softw. 2012;48. doi:10.18637/jss.v048.i02

45. Robin X, Turck N, Hainard A, Tiberti N, Lisacek F, Sanchez J-C, et al. pROC: an open-source package for R and S+ to analyze and compare ROC curves. BMC Bioinformatics. 2011;12: 77. doi:10.1186/1471-2105-12-77

46. Pedersen T. patchwork: The Composer of Plots. 2022. Available: https://CRAN.R-project.org/package=patchwork

47. Jorgensen TD, Pornprasertmanit S, Schoemann AM, Rosseel Y. semTools: Useful tools for structural equation modeling. 2025. Available: https://CRAN.R-project.org/package=semTools

48. Wickham H, Averick M, Bryan J, Chang W, McGowan L, François R, et al. Welcome to the Tidyverse. J Open Source Softw. 2019;4: 1686. doi:10.21105/joss.01686

49. Baron-Cohen S, Cassidy S, Auyeung B, Allison C, Achoukhi M, Robertson S, et al. Attenuation of Typical Sex Differences in 800 Adults with Autism vs. 3,900 Controls. Hu VW, editor. PLoS ONE. 2014;9: e102251. doi:10.1371/journal.pone.0102251

50. Baron-Cohen S. The extreme male brain theory of autism. Trends Cogn Sci. 2002;6: 248–254. doi:10.1016/S1364-6613(02)01904-6

51. Belcher HL, Uglik-Marucha N, Vitoratou S, Ford RM, Morein-Zamir S. Gender bias in autism screening: measurement invariance of different model frameworks of the Autism Spectrum Quotient. BJPsych Open. 2023;9: e173. doi:10.1192/bjo.2023.562

52. Russell-Smith SN, Maybery MT, Bayliss DM. Relationships between autistic-like and schizotypy traits: An analysis using the Autism Spectrum Quotient and Oxford-Liverpool Inventory of Feelings and Experiences. Personal Individ Differ. 2011;51: 128–132. doi:10.1016/j.paid.2011.03.027

53. Lundqvist L-O, Lindner H. Is the Autism-Spectrum Quotient a Valid Measure of Traits Associated with the Autism Spectrum? A Rasch Validation in Adults with and Without Autism Spectrum Disorders. J Autism Dev Disord. 2017;47: 2080–2091. doi:10.1007/s10803-017-3128-y

54. Lau WY-P, Gau SS-F, Chiu Y-N, Wu Y-Y, Chou W-J, Liu S-K, et al. Psychometric properties of the Chinese version of the Autism Spectrum Quotient (AQ). Res Dev Disabil. 2013;34: 294–305. doi:10.1016/j.ridd.2012.08.005

55. Hoekstra RA, Bartels M, Cath DC, Boomsma DI. Factor Structure, Reliability and Criterion Validity of the Autism-Spectrum Quotient (AQ): A Study in Dutch Population and Patient Groups. J Autism Dev Disord. 2008;38: 1555–1566. doi:10.1007/s10803-008-0538-x

56. Zhang L, Sun Y, Chen F, Wu D, Tang J, Han X, et al. Psychometric properties of the Autism-Spectrum Quotient in both clinical and non-clinical samples: Chinese version for mainland China. BMC Psychiatry. 2016;16: 213. doi:10.1186/s12888-016-0915-5

57. Robinson EB, Munir K, Munafò MR, Hughes M, McCormick MC, Koenen KC. Stability of Autistic Traits in the General Population: Further Evidence for a Continuum of Impairment. J Am Acad Child Adolesc Psychiatry. 2011;50: 376–384. doi:10.1016/j.jaac.2011.01.005

58. Fusar-Poli L, Ciancio A, Gabbiadini A, Meo V, Patania F, Rodolico A, et al. Self-Reported Autistic Traits Using the AQ: A Comparison between Individuals with ASD, Psychosis, and Non-Clinical Controls. Brain Sci. 2020;10: 291. doi:10.3390/brainsci10050291

59. Morrier MJ, Ousley OY, Caceres-Gamundi GA, Segall MJ, Cubells JF, Young LJ, et al. Brief Report: Relationship Between ADOS-2, Module 4 Calibrated Severity Scores (CSS) and Social and Non-Social Standardized Assessment Measures in Adult Males with Autism Spectrum Disorder (ASD). J Autism Dev Disord. 2017;47: 4018–4024. doi:10.1007/s10803-017-3293-z

60. Banker SM, Harrington M, Schafer M, Na S, Heflin M, Barkley S, et al. Phenotypic divergence between individuals with self-reported autistic traits and clinically ascertained autism. Nat Ment Health. 2025;3: 286–297. doi:10.1038/s44220-025-00385-8

61. Pisula E, Kawa R, Szostakiewicz Ł, Łucka I, Kawa M, Rynkiewicz A. Autistic Traits in Male and Female Students and Individuals with High Functioning Autism Spectrum Disorders Measured by the Polish Version of the Autism-Spectrum Quotient. Zalla T, editor. PLoS ONE. 2013;8: e75236. doi:10.1371/journal.pone.0075236

62. Gillett G, Leeves L, Patel A, Prisecaru A, Spain D, Happé F. The prevalence of autism spectrum disorder traits and diagnosis in adults and young people with personality disorders: A systematic review. Aust N Z J Psychiatry. 2023;57: 181–196. doi:10.1177/00048674221114603

63. Sizoo BB, Van Den Brink W, Gorissen-van Eenige M, Koeter MW, Van Wijngaarden-Cremers PJM, Van Der Gaag RJ. Using the Autism-Spectrum Quotient to Discriminate Autism Spectrum Disorder from ADHD in Adult Patients With and Without Comorbid Substance Use Disorder. J Autism Dev Disord. 2009;39: 1291–1297. doi:10.1007/s10803-009-0743-2

64. Westwood H, Eisler I, Mandy W, Leppanen J, Treasure J, Tchanturia K. Using the Autism-Spectrum Quotient to Measure Autistic Traits in Anorexia Nervosa: A Systematic Review and Meta-Analysis. J Autism Dev Disord. 2016;46: 964–977. doi:10.1007/s10803-015-2641-0

